# Evaluating Real-World COVID-19 Vaccine Effectiveness Using a Test-Negative Case-Control Design

**DOI:** 10.1101/2022.01.06.22268726

**Authors:** Matthew W Reynolds, Alex Secora, Alice Joules, Lisa Albert, Emma Brinkley, Tom Kwon, Christina Mack, Stephen Toovey, Nancy A. Dreyer

**Affiliations:** IQVIA Real-World Solutions. 201 Broadway, Cambridge, MA 02139 USA; Pegasus Research, Neuschwaendistrasse 6, 6390 Engelberg, Switzerland

## Abstract

It is important to assess the extent to which the real-world effectiveness of marketed vaccines is consistent with that observed in the clinical trials, and to characterize how well vaccines prevent COVID-19 symptoms. We conducted a modified test-negative design (TND) to evaluate the RW effectiveness of three COVID-19 vaccines by leveraging data from an on-going, US community-based registry. Vaccine effectiveness was examined in two ways: considering cases who (1) tested positive for COVID-19 (695 cases, 1,786 controls) and who (2) tested positive with at least one moderate/severe COVID-19 symptom (165 cases, 2,316 controls). Any vaccination (full or partial) was associated with a 95% reduction in the odds of having a positive COVID-19 test [adjusted odds ratio (aOR) = 0.05 (95% confidence interval (CI): 0.04, 0.06)]. Full vaccination was associated with an aOR of 0.03 (95% CI: 0.03, 0.05) while partial vaccination had an aOR of 0.08 (95% CI: 0.06, 0.12). Any vaccination was associated with a 71% reduction in the odds of testing positive and having at least one moderate/severe symptom (aOR=0.29 (95% CI: 0.20, 0.40)). High effectiveness was observed across all three vaccine manufacturers both for prevention of positive COVID-19 test results and prevention of moderate/severe COVID-19 symptoms.

**Clinicaltrials.gov NCT04368065**

## Introduction

Large-scale clinical trials evaluating the currently approved COVID-19 vaccines demonstrated robust efficacy in controlled settings but there are limited data on their effectiveness under real-world conditions, and in particular their effectiveness at preventing more severe symptoms associated with infection.[1-3] Establishing the real-world effectiveness of COVID-19 vaccines may help promote vaccine uptake by allaying concerns about the unprecedented speed at which the vaccines were developed and launched, especially as new variants continue to emerge.[4]

One efficient method of evaluating vaccine effectiveness is the test-negative case control study design (TND),[5-8], often used for studying influenza vaccines where clinical trials may not be ethical or feasible, and formal testing is not routinely conducted.[9-12] They have also recently been used for COVID-19 vaccine effectiveness research since TND studies have been proven to provide reliable estimates of vaccine effectiveness without being subject to confounding by health care seeking behavior.[13-16] These designs differ from traditional case-control study designs in that the controls are distinguished from cases by testing negative for COVID-19, where both cases and controls may have sought testing due to COVID-like symptoms or possible exposure to COVID-19. TND case control studies are useful tools that can inform both regulatory and public health policy, especially since most of the post-marketing United States (US) data on the COVID-19 vaccine effectiveness comes from ecological reporting and observational studies and there is little information on their effectiveness in community-dwelling, non-healthcare affiliated adults.[17-22]

We used a modified version of the TND using direct-to-patient survey data (using community reporting via internet volunteers as opposed to traditional site-based recruitment) to estimate vaccine effectiveness of the Moderna, Pfizer, and Janssen vaccines in the general population, including their effectiveness at preventing moderate to severe COVID-19 symptoms.

## Methods

This study used self-reported data from the COVID-19 Active Research Experience (CARE) registry’s community-based on-line registry, first launched in March, 2020 to study COVID-19 symptoms and severity outside of the hospital setting and to identify what factors, if any, mitigated the risk from COVID-19 (www.helpstopCOVID19.com).[23, 24] The protocol and survey were updated in January 2021 to include information on vaccination, regardless of whether participants had ever contracted COVID-19. Participants are unpaid, recruited primarily via social media and provide informed consent online. Data collection and study conduct followed all ethical principles outlined in the Declaration of Helsinki for the conduct of medical research. This study was approved by an Institutional Review Board and is registered at Clinicaltrials.gov NCT04368065 and EU PAS register EUPAS36240. At enrollment, community-based participants report demographics, COVID-19 test results, medical history, presence and severity of COVID-19-like symptoms on a 4-point scale, as well as use of medications (prescription, non-prescription) and dietary supplements. In January 2021, questions were added regarding COVID-19 vaccination including dates of administrations and manufacturer for each vaccination.[25] Confirmation of vaccination and COVID-19 test results were not independently obtained. COVID-19-related symptoms were assembled from the core FDA list[26] with some additions resulting from common free text write-in responses from the surveys. These symptoms included fever, chills, cough, shortness of breath/difficulty breathing, nasal congestion, sore throat, nausea, diarrhea, fatigue, headache, aches and pains, runny nose, decreased sense of smell, decreased sense of taste, decreased appetite, vomiting, persistent pain or pressure in the chest, trouble waking up after sleeping, anxiety, feeling disoriented or having trouble thinking, depression, and insomnia or trouble sleeping.

We used a modified TND to evaluate the effectiveness of the COVID-19 vaccines. In contrast to a traditional TND that is site-based and prospectively tests all participants at the point of care, this study employs more efficient online recruitment of primarily unpaid volunteers via social media and self-reporting of COVID-19 vaccines, COVID-testing, results, COVID symptoms and severity. While this is a diversion from the traditional approach, these participants have more direct and immediate access to the relevant COVID-testing than to testing for other infectious diseases like influenza. To ensure that the study population had the potential for vaccination, we restricted these analyses to participants who reported a COVID-test result between March 1 through September 16, 2021, a time-period during which vaccines were widely available in the United States.

We approached our analysis in two ways. Our first analysis (COVID-19 case positivity) defines cases as those participants who were tested for any reason and reported a positive COVID-19 test result, and controls as those participants who were tested for any reason and reported a negative COVID-19 result. When participants reported both positive and negative test results in that time interval, we selected the positive result and classified them as a case as of that date. When they reported multiple positive tests, we randomly selected one of the results as the study test result; similarly, if a participant reported multiple negative results, a random result was selected and deemed the study test result. Random selection was used to ensure that there was no bias implemented by always taking earlier or later tests, knowing that access to vaccination increased over time starting early in 2021.

A second analysis incorporates the severity of self-reported COVID-like symptoms. In this analysis, cases were defined as participants who reported a positive COVID-19 test result and also reported at least one moderate or severe COVID-19 symptom within +/- 7 days of that test result. Controls included all participants who reported a negative COVID-19 test result as well as those who reported a positive test result but did not report any moderate/severe symptoms within +/- 7 days of that test result. Symptoms were queried from participants at the time of positive or negative test report; when multiple surveys were available within +/- 7 days of the reported COVID-test, the survey with the largest number of moderate/severe symptoms reported was chosen to assure we identified the highest severity of symptoms reported.

Participants reports were used to determine each person’s vaccination status, which was subsequently characterized as unvaccinated, partially vaccinated, or fully vaccinated at the time of the reported COVID-test result. Unvaccinated participants had not received any vaccine at the time of the COVID-19 test result (Figure 1). Participants were classified as “fully vaccinated” at 14 days post-2nd dose of Moderna or Pfizer vaccine or 14 days after their receipt of the Janssen vaccine. Participants were classified as “partially” vaccinated if they had only received their 1st vaccination of Pfizer or Moderna but not the 2^nd^ dose, or if they had received 2 doses of Pfizer or Moderna vaccine, but were not yet 14 days post-2^nd^ dose, or had received Janssen but were not yet 14 days post-vaccination. Five participants who reported receiving doses from different manufacturers are treated as fully vaccinated using the same time intervals described previously but were excluded from the manufacturer-specific analysis. If participants noted vaccination prior to December 1, 2020 (n=10) or reported vaccination with a COVID-19 vaccine other than Janssen, Pfizer, or Moderna, they were excluded from this study (n=6). Also excluded were those who reported the same date for their first and second vaccine doses for Moderna and Pfizer (n=8). Boosters were largely unavailable at the time these data were collected and are not considered in this study.

**Figure one.**
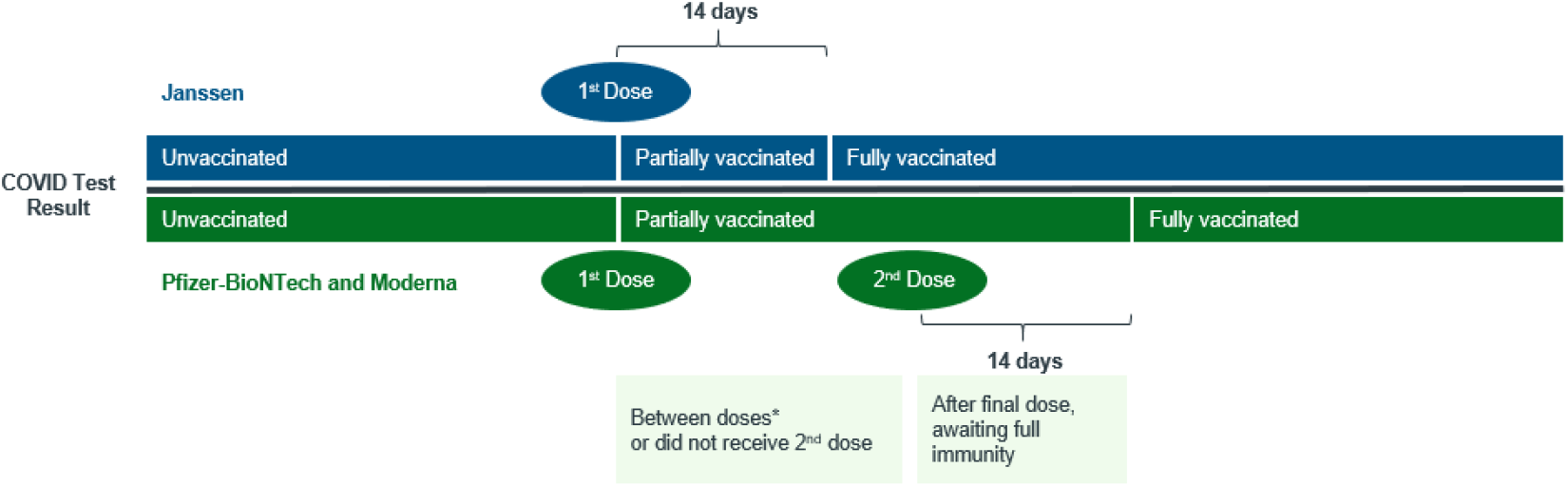
Vaccination status determination at time of COVID-19 testing for single dose and two dose vaccines.

The core analyses aimed to compare the exposure of vaccinated (fully or partially) vs unvaccinated, but also included sensitivity analyses separately for those that were fully vaccinated vs unvaccinated and partially vaccinated vs. unvaccinated. Further, each vaccine manufacture was examined separately for effectiveness using the primary study vaccine exposure definition (fully or partially vaccinated vs unvaccinated).

Access proxy was determined by defining all 50 states and District of Columbia (D.C.) into four categories according to the timing of availability and the administration rate based on vaccine roll-out date for the general population aged 16 and older, identified from each jurisdiction’s government agency website with median being the cut-off in determining high and low. High and low administration rates were determined by vaccine administration per 100,000 population for each state and D.C. on June 1, 2021 according to CDC data[27] for COVID-19 vaccinations in the United States with the median number being the cut off to determine high and low. June 1, 2021 was deemed to be a median date where the majority of the United States population would have an opportunity to access vaccinations. (see Table one).

**Table one:** Access proxy defined as matrix of availability and vaccine administration rate.

|  | **High Vaccination Rate (Admin per 100k on 6/1\*)** | **Low Vaccination Rate (Admin per 100k on 6/1\*)** |
| --- | --- | --- |
| **Early Availability**<br>**(March 10 – April 2)** | CT, NH, CO, MN, IL, OH, AZ, MT, UT, ND, KS<br>(>79,938)<br>Count: 11 | TX, IN, OK, WV, SC, GA, AR, WY, LA, AL, MS<br>(<79,938)<br>Count: 11 |
| **Late Availability**<br>**(April 5 – April 19)** | VT, MA, HI, DC, ME, RI, NM, NJ, MD, NY, WA, PA, VA, CA, DE<br>(>96,910)<br>Count: 15 | OR, WI, IA, NE, SD, FL, MI, AK, KY, NV, NC, MO, TN, ID<br>(<96,910)<br>Count: 14 |

Crude and adjusted odds ratios (95% confidence intervals) were calculated via multivariate logistic regression models to examine vaccine effectiveness at preventing cases of COVID-19. Multivariate odds ratios were adjusted by the following: race (white, other), gender (female, male, other [including male, transgender, not disclosed, other]), age (continuous), gender (female, male, vs. other (including not disclosed, other, transgender), education (some college or less, 4-year college degree, >4 year college degree), ethnicity of Hispanic or Latino (yes, no), access to COVID-19 vaccine proxy (early availability, high vaccination rate; early availability, low vaccination rate; late availability, high vaccination rate; late availability, low vaccination rate; medical conditions including anxiety, autoimmune disorder, blood disorder, cardiovascular disorder, depression, diabetes, hypertension, insomnia or trouble sleeping, kidney disorder, and lung disorder (yes, no).

## Results

### Participant Characteristics

Of the 2,481 participants who were tested and reported vaccination status, 695 individuals reported a positive COVID-19 test result and 1786 individuals reported a negative test result. Among these participants, 1,211 (48.8%) were fully vaccinated, 354 (14.3%) were partially vaccinated, and 916 (36.9%) were unvaccinated (Table two). Most participants (80.2%) reported not having any moderate/severe symptoms; 6.2% having at least one moderate to severe symptom, and 13.6% having two or more such symptoms. Among participants that reported a positive COVID-19 test, 12.5% of cases reported at least one moderate/severe symptom as compared to 3.75% of those reporting a negative test. Study participants had a mean age of 46.4 years (Standard deviation (SD) = 15.3) and were predominately female (82.8%), well educated (61.9% ≥ 4-year college), and white (87.1%). Both cases and controls were similar with regard to age (mean age: 46.7(SD= 15.3) vs. 46.3(SD = 15.0)), gender (female 80.7% vs 83.7%), and race (white: 89.5% vs 86.2%), but the case population (52.5% ≥ 4-year college) reported slightly higher education than controls (61.9% ≥ 4-year college). A notable proportion of the study participants reported having anxiety (38.5%), depression (32.5%), insomnia or trouble sleeping (29.8%), and/or hypertension (22.6%) at their baseline survey, which were comparable across both cases and controls.

**Table two.** Participant characteristics.

|  | Cases<br>COVID-19 test positive<br>N=695 | Controls<br>COVID-19 test negative<br>N=1786 | Cases<br>COVID-19 test<br>positive with >1<br>moderate to severe COVID<br>symptoms<br>N=165 | Controls<br>COVID-19 test positive with <1<br>moderate to severe COVID<br>symptoms & COVID-19<br>test negative<br>N=2316 | Total<br>N=2481 |
| --- | --- | --- | --- | --- | --- |
| Vaccination Status (N) | 695 | 1786 | 165 | 2316 | 2481 |
| Fully Vaccinated | 76 (10.9) | 1135 (63.6) | 42 (25.5) | 1199 (51.8) | 1211 (48.8) |
| Partially Vaccinated | 48 (6.9) | 306 (17.1) | 15 (9.1) | 348 (15.0) | 354 (14.3) |
| Unvaccinated | 571 (82.2) | 345 (19.3) | 108 (65.5) | 769 (33.2) | 916 (36.9) |
| Number of Moderate/Severe<br>Symptoms |  |  |  |  |  |
| None | 431 (62.0) | 1559 (87.3) | 0 (0) | 1991 (85.9) | 1990 (80.2) |
| One | 87 (12.5) | 67 (3.75) | 54 (32.7) | 99 (4.3) | 154 (6.2) |
| Two or more | 177 (25.5) | 160 (8.9) | 111 (67.3) | 226 (9.8) | 337 (13.6) |
| Age in years (N) | 695 | 1786 | 165 | 2316 | 2481 |
| Mean (SD) | 46.7 (15.3) | 46.3 (15.0) | 46.7 (15.2) | 46.3 (15.1) | 46.4 (15.1) |
| Median [IQR] | 46 (34 – 60) | 45 (34 – 59) | 46 (34 – 59) | 45 (34 – 59) | 45 (34 – 59) |
| Age Group (years) | 695 | 1786 | 19 (11.5) | 322 (13.9) | 2481 |
| 18-29 | 93 (13.4) | 248 (13.9) | 35 (21.2) | 573 (24.7) | 341 (13.74) |
| 30-39 | 164 (23.6) | 444 (24.8) | 27 (16.4) | 433 (18.7) | 608 (24.5) |
| 40-49 | 123 (17.7) | 337 (18.9) | 45 (27.3) | 416 (17.9) | 460 (18.5) |
| 50-59 | 139 (20.0) | 322 (18.0) | 39 (23.6) | 572 (24.7) | 461 (18.6) |
| >=60 | 176 (25.3) | 435 (24.4) | 19 (11.5) | 322 (13.9) | 611 (24.6) |
| Gender | 695 | 1786 | 165 | 2316 | 2481 |
| Female | 561 (80.7) | 1495 (83.7) | 135 (81.8) | 1921 (82.9) | 2056 (82.8) |
| Male | 118 (16.9) | 244 (13.7) | 25 (15.2) | 337 (14.6) | 362 (14.6) |
| Transgender | 7 (1.0) | 26 (1.5) | 2 (1.2) | 31 (1.3) | 33 (1.3) |
| Other | 9 (1.3) | 20 (1.5) | 3 (1.8) | 26 (1.1) | 29 (1.2) |
| Not Disclosed | 0 (0) | 1 (0.1) | 0 (0) | 1 (0.04) | 1 (0.04) |
| Education | 695 | 1784 | 165 | 2314 | 2479 |
| Some college or less | 330 (47.5) | 614 (34.4) | 88 (53.3) | 856 (37.0) | 944 (38.1) |
| 4-yr college degree | 157 (22.6) | 491 (27.5) | 36 (21.8) | 612 (26.4) | 648 (26.1) |
| >4yr college degree | 208 (29.9) | 679 (38.1) | 41 (24.9) | 846 (36.5) | 887 (35.8) |
| Race | 692 | 1783 | 1 (0.6) | 53 (2.3) | 2481 |
| Asian | 5 (0.7) | 49 (2.8) | 2 (1.2) | 81 (3.5) | 54 (2.2) |
| Black or African American | 24 (3.5) | 59 (3.3) | 143 (86.7) | 2013 (86.9) | 83 (3.4) |
| White | 619 (89.5) | 1537 (86.2) | 18 (10.9) | 164 (7.1) | 2156 (87.1) |
| Other** | 44 (6.4) | 138 (7.7) | 1 (0.6) | 5 (0.2) | 182 (7.4) |
| Ethnicity |  |  |  |  |  |
| Hispanic or Latino | 73 (10.6) | 154 (8.7) | 14 (8.5) | 213 (9.2) | 227 (9.2) |
| Medical Conditions |  |  |  |  |  |
| Anxiety | 289 (41.5) | 666 (37.3) | 84 (50.9) | 871 (37.6) | 955 (38.5) |
| Autoimmune disease | 90 (13.0) | 214 (11.9) | 27 (16.3) | 277 (11.9) | 304 (12.3) |
| Blood disorder | 30 (4.3) | 36 (2.0) | 54 (2.3) | 12 (7.3) | 66 (2.7) |
| Cardiovascular disease | 39 (5.6) | 100 (5.6) | 14 (8.5) | 125 (5.4) | 139 (5.6) |
| Depression | 239 (34.4) | 567 (31.8) | 69 (41.8) | 737 (31.8) | 806 (32.5) |
| Diabetes | 65 (9.4) | 151 (8.5) | 15 (9.1) | 201 (8.7) | 216 (8.7) |
| Hypertension | 158 (22.7) | 402 (22.5) | 40 (24.2) | 520 (22.5) | 560 (22.6) |
| Insomnia or trouble sleeping | 225 (32.4) | 515 (28.8) | 72 (43.6) | 668 (28.8) | 740 (29.8) |
| Kidney disease | 21 (3.02) | 53 (3.0) | 6 (3.6) | 68 (2.9) | 74 (3.0) |
| Lung disease | 63 (9.06) | 204 (11.4) | 21 (12.7) | 246 (10.6) | 267 (10.7) |
| Organ transplant | 2 (0.3) | 3 (0.2) | 1 (0.6) | 4 (0.2) | 5 (0.2) |
| On Treatment for Cancer? | 692 | 1777 |  |  | 2469 |
| Yes | 10 (1.5) | 22 (1.2) | 5 (3.0) | 27 (1.2) | 32 (1.3) |

\* The race category of other includes: 13 American Indians or Alaskans, 3 Hawaiian or Pacific Islander, 77 who selected multiple races and 89 who selected 'other'.

### Test Negative Design Analysis

For the first analysis, which focused on evaluating COVID-19 vaccine effectiveness at preventing a COVID-19 positive test, 124 cases (17.8% of 695) reported being fully or partially vaccinated at the time of their positive COVID-19 test, while 1441 controls (80.6% of 1,786) reported being fully or partially vaccinated at the time of their negative COVID-19 test (Table two). This resulted in an unadjusted odds ratio (aOR) of 0.05 (95% CI: 0.04, 0.07) and adjusted odds ratio of 0.05 (95% CI: 0.04, 0.06), indicating that being vaccinated was associated with a 95% reduction in the odds of having a positive COVID-19 test.

The second analysis focused on evaluating the effectiveness of preventing at least one moderate/severe COVID-19 symptom in COVID-19 positive cases and included 165 cases and 2,316 controls (Table three). Fifty-seven cases reported being fully or partially vaccinated at the time of their positive COVID-19 test with at least one moderate/severe symptom, while 1547 control patients reported being fully or partially vaccinated at the time of their negative COVID-19 test with at least on moderate/severe symptom. This resulted in an unadjusted odds ratio of 0.26 (95% CI: 0.19, 0.37) and adjusted odds ratio of 0.29 (95% CI: 0.20, 0.40), indicating a 71% reduction in the odds of having a positive COVID-19 test and at least one moderate/severe symptom.

**Table three.** Multivariate model for vaccine effectiveness at preventing cases of COVID-19.

| Covariates | Odds of having a COVID-19 positive test | Odds of having a COVID-19 positive test with $\geq 1$ moderate/severe symptom) |
| --- | --- | --- |
|  | Adjusted OR (95% CI) | Adjusted OR (95% CI) |
| Vaccination Status |  |  |
| Unvaccinated | (ref) | (ref) |
| Fully or Partially Vaccinated | 0.05 (0.04, 0.06) | 0.29 (0.20, 0.40) |
| Anxiety | 1.27 (0.96, 1.69) | 1.46 (0.97, 2.21) |
| Autoimmune Disorder | 1.22 (0.86, 1.74) | 1.15 (0.72, 1.85) |
| Blood Disorder | 3.01 (1.56, 5.83) | 2.60 (1.28, 5.28) |
| Cardiovascular Disorder | 0.89 (0.53, 1.50) | 1.47 (0.76, 2.81) |
| Depression | 1.00 (0.74, 1.33) | 1.07 (0.71, 1.61) |
| Diabetes | 1.23 (0.81, 1.86) | 0.87 (0.48, 1.58) |
| Hypertension | 0.89 (0.66, 1.20) | 0.85 (0.55, 1.32) |
| Insomnia or trouble sleeping | 1.09 (0.84, 1.42) | 1.57 (1.09, 2.25) |
| Kidney Disorder | 1.19 (0.60, 2.36) | 0.95 (0.38, 2.42) |
| Lung Disorder | 0.55 (0.37, 0.81) | 0.90 (0.54, 1.52) |
| Education |  |  |
| Some college or less | (ref) | (ref) |
| 4 years of college | 0.73 (0.55, 0.97) | 0.71 (0.46, 1.08) |
| More than 4 years of college | 0.97 (0.74, 1.27) | 0.68 (0.45, 1.02) |
| Race |  |  |
| Other | (ref) | (ref) |
| White | 2.33 (1.64, 3.32) | 1.09 (0.65, 1.82) |
| Age (per 1 year increment) | 1.00 (0.99, 1.01) | 1.01 (0.99, 1.02) |
| Gender |  |  |
| Female | (ref) | (ref) |
| Male | 1.50 (1.10, 2.06) | 1.26 (0.79, 2.01) |
| Other | 1.35 (0.63, 2.91) | 1.29 (0.48, 3.47) |
| Ethnicity (Hispanic Latino vs. Not) |  |  |
| Not Hispanic | (ref) | (ref) |
| Hispanic | 0.99 (0.68, 1.46) | 0.84 (0.45, 1.54) |
| Vaccination Access (Proxy) |  |  |
| Early availability, high vaccination rate | (ref) | (ref) |
| Early availability, low vaccination rate | 0.87 (0.60, 1.26) | 1.24 (0.74, 2.09) |
| Late availability, high vaccination rate | 0.63 (0.46, 0.86) | 0.72 (0.44, 1.16) |
| Late availability, low vaccination rate | 1.15 (0.83, 1.60) | 1.26 (0.79, 2.02) |

Note: If a referent category is not listed, then the comparison is Yes vs. No (referent group).

When examining the effects of covariates on reporting a positive test for COVID-19, male gender (OR 1.50; 95% CI: 1.10, 2.06), white race (OR 2.33; 95% CI: 1.64, 3.32), reporting a blood disorder, (including a history of blood clots, sickle cell disease, thalassemia, thrombocytopenia, or other blood disease), (OR 3.01: 95% CI: 1.56, 5.83), and reporting a lung disorder (OR 0.55, 95% CI: 0.37, 0.81) were significant predictors in the multivariate model. While not statistically significant, low vaccination rates in the state of the participant, regardless of timing of vaccine availability were suggestive of increased risk of COVID-19 positive testing (early availability/low access = OR 1.24; 95% CI: 0.74, 2.09; and late availability/low access = OR 1.26; 95% CI 0.79, 2.02).

When examining the effectiveness of full vaccination (versus being unvaccinated) and partial vaccination (at least one dose, but not having achieved full potential immunity) versus being unvaccinated, full vaccination was more protective (aOR=0.03, 95% CI: 0.03, 0.05) when compared to partial vaccination (versus being unvaccinated) (aOR=0.08, 95% CI: 0.06, 0.12) (Table four). In the symptom-based analysis, fully vaccination (aOR=0.28, 95% CI: 0.19, 0.41)) resulted in slightly better effectiveness than those with partial vaccination (aOR=0.29, 95% CI: 0.16, 0.53).

**Table four.** Test negative analysis (odds of a positive COVID-19 test or a positive COVID-19 test and at least one moderate to severe symptom by vaccination status.

| Test negative analysis based on COVID-19 test results | Vaccination Status | Cases COVID-19 test positive | Controls COVID-19 test negative | OR (95% CI) | aOR (95% CI) |
| --- | --- | --- | --- | --- | --- |
|  | Fully Vaccinated | 76 | 1135 | 0.04 (0.03, 0.05) | 0.03 (0.03, 0.05) |
|  | Unvaccinated | 571 | 345 |  |  |
|  | Partially Vaccinated | 48 | 306 | 0.10 (0.07, 0.13) | 0.08 (0.06, 0.12) |
|  | Unvaccinated | 571 | 345 |  |  |
| Test negative analysis based on COVID-19 test results and requiring at least one moderate to severe symptom | Vaccination Status | Cases COVID-19 test positive with >1 moderate to severe COVID symptoms | Controls COVID-19 test positive with <1 moderate to severe COVID symptoms & COVID-19 test negative | OR (95% CI) | aOR (95% CI) |
|  | Fully vaccinated | 42 | 1199 | 0.25 (0.17, 0.36) | 0.28 (0.19, 0.41) |
|  | Unvaccinated | 108 | 769 |  |  |
|  | Partially vaccinated | 15 | 348 | 0.31 (0.18, 0.53) | 0.29 (0.16, 0.53) |
|  | Unvaccinated | 108 | 769 |  |  |

A manufacturer-specific test negative design analysis was also conducted via both case definitions. When examining COVID-19 test positivity alone, vaccine effectiveness results were similar across manufacturers, conveying significant protection against COVID-19 infection (Table five and Figure two). Moderna had the highest effectiveness via Odds Ratios (97%, aOR=0.03, 95% CI: 0.02, 0.04), followed closely by Janssen at 96% (aOR=0.04, 95% CI: 0.02, 0.09), and Pfizer BioNTech at 94% (aOR=0.06, 95% CI: 0.04, 0.07). When examining by the symptom severity case definition, Moderna showed the best results at preventing symptomatic cases with an 86% effectiveness (aOR=0.14, 95% CI: 0.07, 0.28), followed by Janssen at 65% (aOR=0.35, 95% CI: 0.11, 1.15) and Pfizer at 62% (aOR=0.38, 95% CI: 0.24, 0.58).

**Table five.**
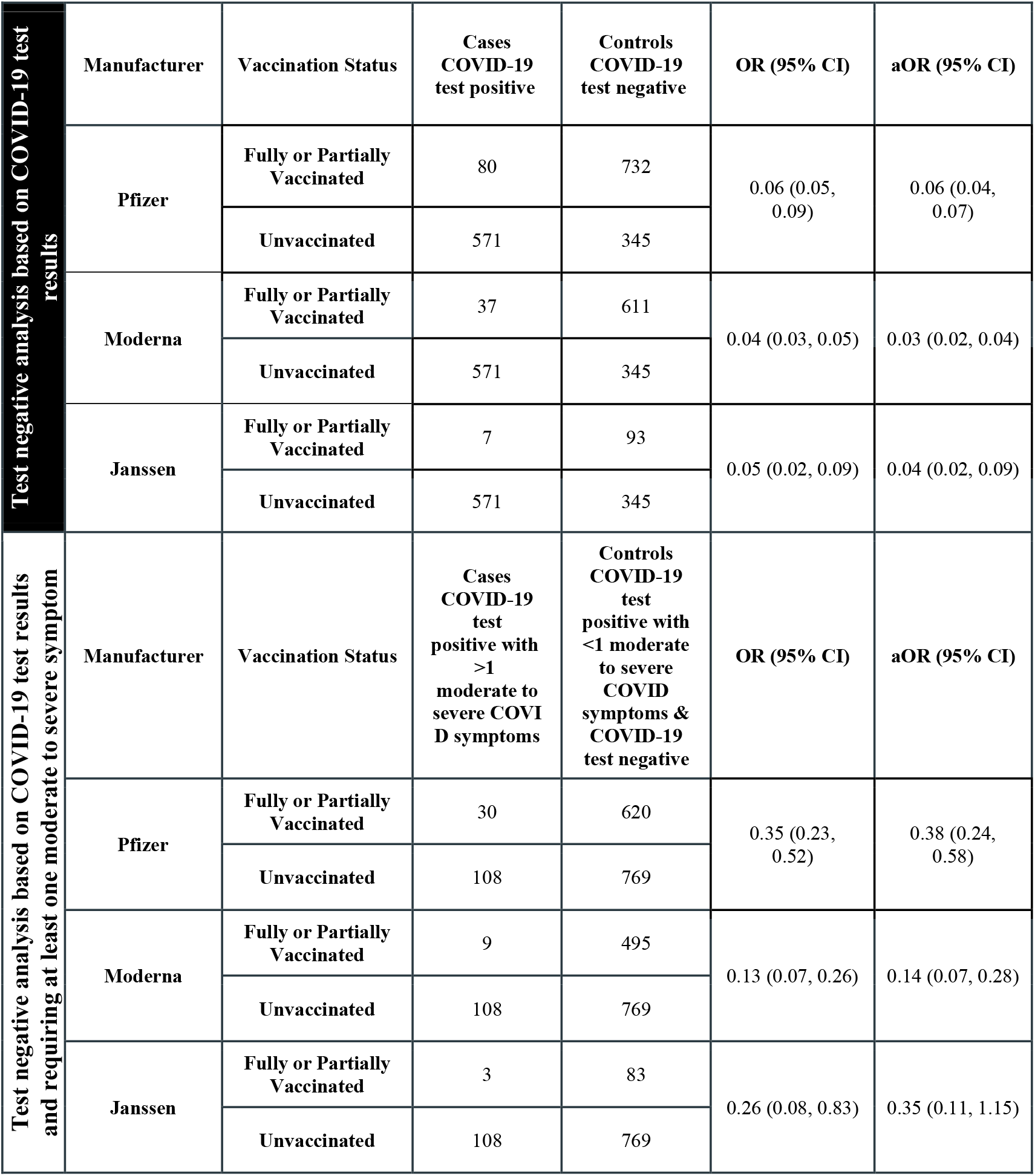
Vaccine effectiveness at preventing cases of COVID-19 by vaccine manufacturer and vaccination status for both test negative analysis results.

**Figure two.**
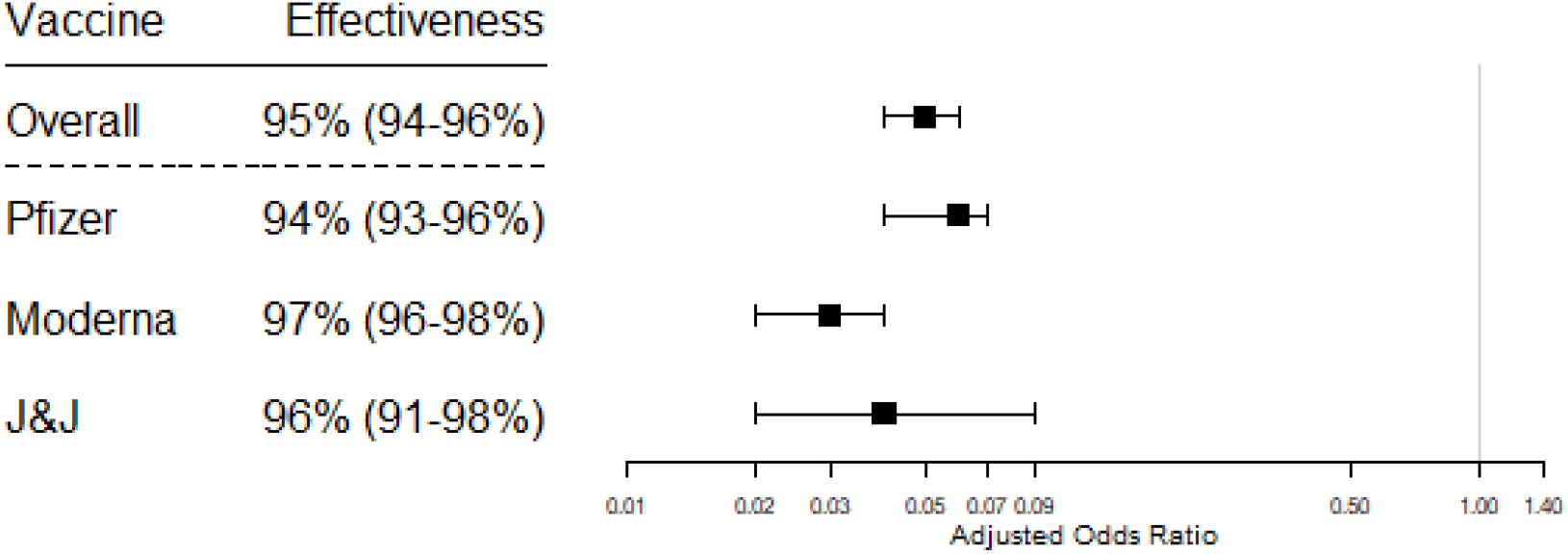
Vaccine effectiveness of preventing cases of COVID-19 among those with at least one vaccination vs unvaccinated, overall and by manufacturer (primary analysis based on COVID-19 test results)

**Figure three.**
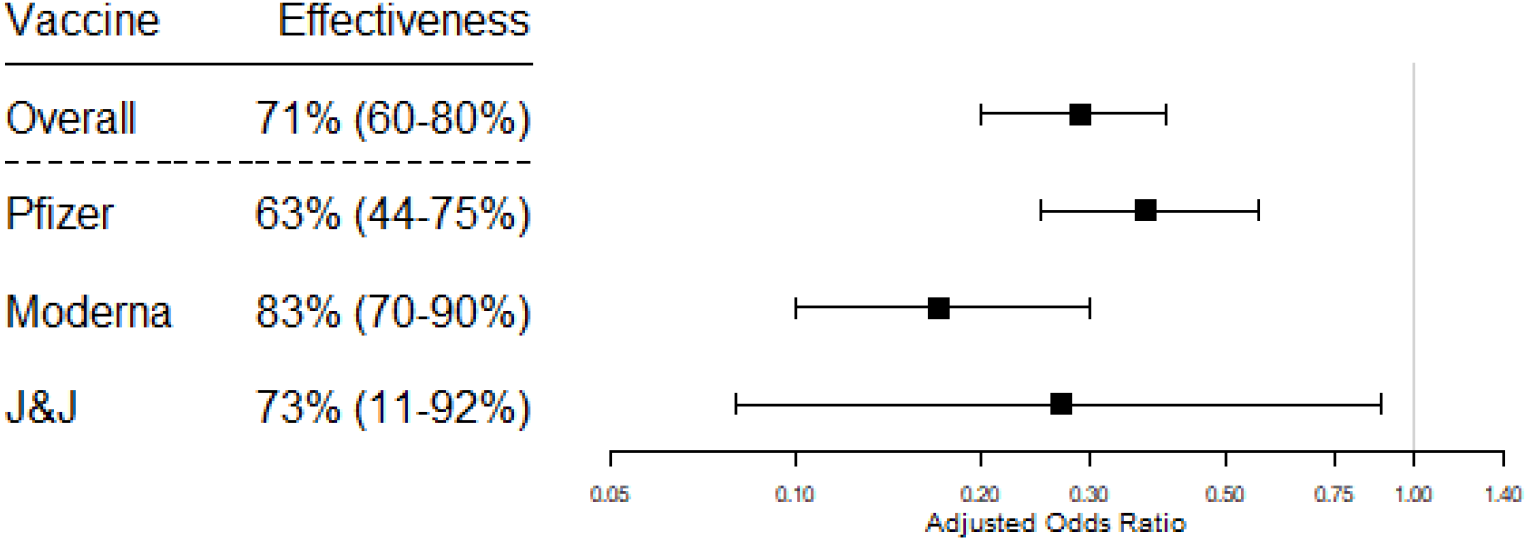
Vaccine effectiveness of preventing cases of COVID-19 among those with at least one vaccination vs unvaccinated, overall and by manufacturer (secondary analysis based on test results and requiring at least one moderate to severe symptom)

## Discussion

While the COVID-19 vaccines were tested through clinical trials establishing their safety and efficacy, the expedited clinical development coupled with immediate widespread use has brought scrutiny from the public as to whether the findings would be replicated in the real-world. Here we show that COVID-19 vaccines manufactured by Pfizer BioNTech, Janssen, and Moderna demonstrated consistent and meaningful real-world effectiveness. The results from this community-based registry of US adults are consistent with what has been reported in randomized controlled trials[1-3] as well as other published TND studies from both ambulatory and inpatient settings.[13-16] We observed a 95% reduction in odds of a COVID-19 positive test in vaccinated participants, with 94%, 96%, and 97% reduction in odds for Pfizer BioNTech, Janssen, and Moderna, respectively. With respect to vaccine effectiveness at mitigating disease severity, we observed a 74% reduction in risk of moderate-to-severe cases overall. Our results suggest greater protection for those who were fully vaccinated as compared to those partially vaccinated, although the results still show clear protection for patients who only received one mRNA vaccine dose and/or had not completed the full immunity period (i.e., 14 days post-final vaccination). These findings contribute to the evidence supporting COVID-19 vaccine effectiveness at mitigating infection in the community overall and also reducing COVID-19 severity among breakthrough infections.

Since their emergency use authorizations, several cohort studies among healthcare workers and among older adults have shown effectiveness of the mRNA vaccines (i.e., Pfizer and Moderna) consistent with the 94-95% efficacy observed Phase III studies.[17, 19, 22] TND studies have also been conducted in similar high-risk groups from the U.S., U.K., and Canada, with consistent findings.[13-16] At the same time, there are still limited data on vaccine effectiveness among community-dwelling adults across age groups, particularly for those receiving the Janssen vaccine. Our results generally align with other cohort and case-control studies in terms of further demonstration of vaccine effectiveness in the general population. Of note, our findings with respect to mRNA vaccine effectiveness appear very close to what was observed in the Phase II studies other than for the Janssen vaccine (96.0% effective in this study versus 66.3% in the clinical trial).

Our study also contributes to the broader understanding of vaccine effectiveness at mitigating moderate to severe COVID-19 disease symptoms. While we observed the strongest effectiveness in the context of preventing a positive test, these COVID-19 vaccines had substantial (∼70%) effectiveness in preventing one or more moderate to severe symptoms, noting that the severity of COVID-19 symptoms here requires self-reporting, which may be difficult from a hospital bed or among very sick patients, and had no requirement for or link to healthcare utilization (this study only included 21 participants that reported any hospitalizations during the study period). Our findings show that COVID-19 vaccines protect against moderate-to severe symptoms that did not necessarily require hospitalization or even interactions with the healthcare system.

While vaccination status was clearly the most important variable in the multivariate models at explaining the risk of a positive COVID-19 test, there were several other variables that were significant and may provide insight into the bigger picture of COVID-19 risk and vaccine effectiveness. Participants who reported a blood disorder in their baseline medical history were associated with a 3-fold risk of testing positive for COVID-19. This blood disorder variable is broad, with our 66 respondents having been prompted by examples that included blood clots, sickle cell disease, thalassemia, thrombocytopenia, or other blood disease. Our finding in this heterogenous group requires some explanation. While it is known that COVID-19 infection may be pro-thrombotic, and this may potentially exacerbate pro-thrombotic blood disorders, our broad and heterogenous ‘blood disorder’ group may also have included conditions associated with immunosuppression, or the use of immunosuppressive medications, in turn resulting in decreased viral clearance and more severe disease. This result may highlight another potential at-risk population for increased risk of the COVID-19 infection, and merits further investigation to better define the heamatological conditions associated with greater risk.Anaemia has been previously suggested to be an independent risk factor for Covid-19 related mortality, and it would be very likely that our group also included anaemic patients, but their exact contribution to the rather large effect observed is uncertain in our data set. [28, 29] Also, being Caucasian was associated here with a more than a twofold increased risk of testing COVID-19 positive, but no elevated risk of testing positive and being symptomatic; however, we suspect that white race may actually be serving here as a proxy for access to COVID-19 testing rather than having a direct causal relationship.

In contrast, participants who reported lung disorders were significantly less likely to test positive for COVID-19 (aOR = 0.55; 95% CI: 0.37, 0.81). It is possible that there may be a common drug/treatment in this group of participants that maybe be COVID-19 protective, but it also makes sense that those at highest risk of severe COVID-19 outcomes may embark on other effective methods at preventing COVID-19 (such as social isolation, etc.), that are not explicitly captured as stand-alone variables in our model. These social variables also play an important role in explaining COVID-19 transmission outside of vaccination, and they should be considered when possible in other studies.

It was also seen that while early availability of the vaccine in the participants’ home states did not have a notable impact, the level of vaccinations in those states did seem to suggest some association with testing positive. Low vaccination rates in the state of the participant, regardless of timing of vaccine availability, show a modest increased risk of COVID-19 positive testing (early availability/low access = OR 1.24; 95% CI: 0.74, 2.09; and late availability/low access = OR 1.26; 95% CI 0.79, 2.02). This suggests that independent of the study participant’s personal vaccination status, if the state in which they live was in the lower half of vaccination rates, their risk of being a COVID-19 positive case was approximately 25% higher. This clearly suggests that increasing the vaccination rate in the population has a beneficial effect at preventing COVID-19 transmission.

While the data are reassuring on the effectiveness of the current COVID-19 vaccines, those vaccinated may not be as strongly protected against emerging variants such as delta and omicron. Coupled with the known possibility of waning immunity from vaccination, additional booster doses are now recommended, particularly for higher risk groups. While cohort studies can be limited when timely data are needed to inform public health measures, the efficiency of a TND or modified TND approach with an unbiased selection of cases and controls are likely one of the best options to study the vaccines in the post-market setting.

Although CARE data rely on self-reported information subject to some information biases, these real-world data have detailed information on participant experiences with respect to COVID-19 and the vaccines. Misclassification of vaccination status can bias TND studies, but is unlikely here since CARE participants are likely to know whether they have been vaccinated and were encouraged to consult their vaccination cards when reporting manufacturer, lot and dates. Moreover, person-generated data like this allows for a comprehensive capture of all relevant patient-level clinical and nonclinical data which are often not available in larger real world data sources. At the same time, it is not always clear whether participants were tested for COVID-19 because they were symptomatic or exposed to someone who tested positive, or if it was mandated for some other reason. It is possible that vaccinated individuals are more likely to take a COVID-19 test out of caution, whereas unvaccinated individuals are not concerned about COVID-19, driving their unvaccinated status., which could bias TND studies if testing practices were differentially affected by vaccination status. While the study was conducted in a period not affected by the more transmissible Delta and Omicron variants, we chose a period that reflects the circulating virus during the Phase III clinical trials to assess consistency of results. This approach, design, and data collection could also be implemented and/or re-directed in an efficient and effective manner for new and emerging COVID-19 vaccine and treatment effectiveness questions.

## Data Availability

Due to data privacy and security regulations the researchers are not able to share participant level data.

## Ethics approval and informed consent

This study was approved by Institutional Review Board and registered at Clinicaltrials.gov NCT04368065 and EU PAS register EUPAS36240. All participants provided informed consent online.

## Consent for publication

Consent for publication of research finding was provided online.

## References

(1) Baden LR, et al. Efficacy and Safety of the mRNA-1273 SARS-CoV-2 Vaccine. N Engl J Med 2021; 384(5): 403–416.

(2) Polack FP, et al. Safety and Efficacy of the BNT162b2 mRNA Covid-19 Vaccine. N Engl J Med 2020; 383(27): 2603–2615.

(3) Sadoff J, et al. Safety and Efficacy of Single-Dose Ad26.COV2.S Vaccine against Covid-19. N Engl J Med 2021; 384(23): 2187–2201.

(4) Patel MM, Jackson ML, Ferdinands J. Postlicensure Evaluation of COVID-19 Vaccines. JAMA 2020; 324(19): 1939–1940.

(5) Chua H, et al. The Use of Test-negative Controls to Monitor Vaccine Effectiveness: A Systematic Review of Methodology. Epidemiology 2020; 31(1): 43–64.

(6) Dean NE, Hogan JW, Schnitzer ME. Covid-19 Vaccine Effectiveness and the Test-Negative Design. N Engl J Med 2021; 385(15): 1431–1433.

(7) Organization WH. Evaluation of COVID-19 vaccine effectiveness: interim guidance, 17 March 2021: World Health Organization; 2021.

(8) Organization WH. Estimating COVID-19 vaccine effectiveness against severe acute respiratory infections (SARI) hospitalisations associated with laboratory-confirmed SARS-CoV-2: an evaluation using the test-negative design: guidance document: World Health Organization. Regional Office for Europe; 2021.

(9) Feng S, et al. Estimating Influenza Vaccine Effectiveness With the Test-Negative Design Using Alternative Control Groups: A Systematic Review and Meta-Analysis. Am J Epidemiol 2018; 187(2): 389–397.

(10) Fukushima W, Hirota Y. Basic principles of test-negative design in evaluating influenza vaccine effectiveness. Vaccine 2017; 35(36): 4796–4800.

(11) Shi M, et al. A comparison of the test-negative and the traditional case-control study designs for estimation of influenza vaccine effectiveness under nonrandom vaccination. BMC Infect Dis 2017; 17(1): 757.

(12) Vasileiou E, et al. Seasonal Influenza Vaccine Effectiveness in People With Asthma: A National Test-Negative Design Case-Control Study. Clin Infect Dis 2020; 71(7): e94–e104.

(13) Lopez Bernal J, et al. Effectiveness of the Pfizer-BioNTech and Oxford-AstraZeneca vaccines on covid-19 related symptoms, hospital admissions, and mortality in older adults in England: test negative case-control study. BMJ 2021; 373: n1088.

(14) Chung H, et al. Effectiveness of BNT162b2 and mRNA-1273 covid-19 vaccines against symptomatic SARS-CoV-2 infection and severe covid-19 outcomes in Ontario, Canada: test negative design study. BMJ 2021; 374: n1943.

(15) Thompson MG, et al. Effectiveness of Covid-19 Vaccines in Ambulatory and Inpatient Care Settings. N Engl J Med 2021; 385(15): 1355–1371.

(16) Pilishvili T, et al. Interim Estimates of Vaccine Effectiveness of Pfizer-BioNTech and Moderna COVID-19 Vaccines Among Health Care Personnel -33 U.S. Sites, January-March 2021. MMWR Morb Mortal Wkly Rep 2021; 70(20): 753–758.

(17) Fowlkes A, et al. Effectiveness of COVID-19 Vaccines in Preventing SARS-CoV-2 Infection Among Frontline Workers Before and During B.1.617.2 (Delta) Variant Predominance - Eight U.S. Locations, December 2020-August 2021. MMWR Morb Mortal Wkly Rep 2021; 70(34): 1167–1169.

(18) Kow CS, Hasan SS. Real-world effectiveness of BNT162b2 mRNA vaccine: a meta-analysis of large observational studies. Inflammopharmacology 2021; 29(4): 1075–1090.

(19) Swift MD, et al. Effectiveness of Messenger RNA Coronavirus Disease 2019 (COVID-19) Vaccines Against Severe Acute Respiratory Syndrome Coronavirus 2 (SARS-CoV-2) Infection in a Cohort of Healthcare Personnel. Clin Infect Dis 2021; 73(6): e1376–e1379.

(20) Britton A, et al. Effectiveness of the Pfizer-BioNTech COVID-19 Vaccine Among Residents of Two Skilled Nursing Facilities Experiencing COVID-19 Outbreaks - Connecticut, December 2020-February 2021. MMWR Morb Mortal Wkly Rep 2021; 70(11): 396–401.

(21) Tenforde MW, et al. Effectiveness of Pfizer-BioNTech and Moderna Vaccines Against COVID-19 Among Hospitalized Adults Aged >/=65 Years - United States, January-March 2021. MMWR Morb Mortal Wkly Rep 2021; 70(18): 674–679.

(22) Thompson MG, et al. Interim Estimates of Vaccine Effectiveness of BNT162b2 and mRNA-1273 COVID-19 Vaccines in Preventing SARS-CoV-2 Infection Among Health Care Personnel, First Responders, and Other Essential and Frontline Workers - Eight U.S. Locations, December 2020-March 2021. MMWR Morb Mortal Wkly Rep 2021; 70(13): 495–500.

(23) Dreyer NA, et al. Self-reported symptoms from exposure to Covid-19 provide support to clinical diagnosis, triage and prognosis: An exploratory analysis. Travel Med Infect Dis 2020; 38: 101909.

(24) Dreyer N, et al. Identification of a Vulnerable Group for Post-Acute Sequelae of SARS-CoV-2 (PASC): People with Autoimmune Diseases Recover More Slowly from COVID-19. Int J Gen Med 2021; 14: 3941–3949.

(25) Dreyer NA, et al. How frequent are acute reactions to COVID-19 vaccination and who is at riskã medRxiv 2021: 2021.2010.2014.21265010.

(26) United States Food and Drug Administration. Assessing COVID-19-Related Symptoms in Outpatient Adult and Adolescent Subjects in Clinical Trials of Drugs and Biological Products for COVID-19 Prevention or Treatment. In, 2020.

(27) Center for Disease Control and Prevention. COVID-19 Vaccinations in the United States. In, 2021.

(28) Roy NBA, et al. Protecting vulnerable patients with inherited anaemias from unnecessary death during the COVID-19 pandemic. Br J Haematol 2020; 189(4): 635–639.

(29) Tremblay D, et al. Mild anemia as a single independent predictor of mortality in patients with COVID-19. EJHaem 2021.

